# Inequalities in colorectal cancer screening uptake in Wales: an examination of the impact of the temporary suspension of the screening programme during the COVID-19 pandemic

**DOI:** 10.1101/2022.08.08.22278493

**Authors:** Diana Bright, Sharon Hillier, Jiao Song, Dyfed W. Huws, Giles Greene, Karen Hodgson, Ashley Akbari, Rowena Griffiths, Alisha R. Davies, Ardiana Gjini

## Abstract

**Background:** Response to the early stages of the COVID-19 pandemic resulted in the temporary disruption of cancer screening in the UK, and strong public messaging to stay safe and to protect NHS capacity. Following reintroduction in services, we explored the impact on inequalities in uptake of the Bowel Screening Wales (BSW) programme to identify groups who may benefit from tailored interventions.

**Methods:** Records within the BSW were linked to electronic health records (EHR) and administrative data within the Secured Anonymised Information Linkage (SAIL) Databank. Ethnic group was obtained from a linked data method available within SAIL. We examined uptake for the first 3 months of invitations (August to October) following the reintroduction of BSW programme in 2020, compared to the same period in the preceding 3 years. Uptake was measured across a 6 month follow-up period. Logistic models were conducted to analyse variations in uptake by sex, age group, income deprivation quintile, urban/rural location, ethnic group, and clinically extremely vulnerable (CEV) status in each period; and to compare uptake within sociodemographic groups between different periods.

**Results:** Uptake during August to October 2020 (period 2020/21; 60.4%) declined compared to the same period in 2019/20 (62.7%) but remained above the 60% Welsh standard. Variation by sex, age, income deprivation, and ethnic groups was observed in all periods studied. Compared to the pre-pandemic period in 2019/20, uptake declined for most demographic groups, except for older individuals (70-74 years) and those in the most income deprived group. Uptake continues to be lower in males, younger individuals, people living in the most income deprived areas and those of Asian and unknown ethnic backgrounds.

**Conclusions:** Our findings are encouraging with overall uptake achieving the 60% Welsh standard during the first three months after the programme restarted in 2020 despite the disruption. Inequalities did not worsen after the programme resumed activities but variations in CRC screening in Wales associated with sex, age, deprivation and ethnicity remain. This needs to be considered in targeting strategies to improve uptake and informed choice in CRC screening to avoid exacerbating disparities in CRC outcomes as screening services recover from the pandemic.

## Introduction

Colorectal cancer (CRC) is estimated to be the third most common cancer and the second leading cause of cancer death globally. In Wales, over 900 people die from the disease every year [1, 2]. Population-level screening programmes have been shown to reduce long-term CRC cancer mortality by up to 27% [3, 4]. However, the success of CRC screening largely depends on uptake among the invited population [5]. The Bowel Wales Screening (BSW) programme has invited eligible population (60-74 year olds) every 2-years, and since October 2021, the programme has been expanded to include 58-74 years olds. In 2019, the BSW programme introduced a more accurate test, the easier-to-use liquid faecal immunochemical test (FIT) instead of the guaiac-based faecal occult blood test (gFOBT) [6], as a strategy to reduce inequity of uptake as this is simpler to use and more acceptable (one stool sample instead of the three required for gFOBT).

Previous studies have shown that in universal population-based disease prevention and promotion programmes, significant social inequalities exist by socioeconomic status, sex, and age [7-9]. In addition, several international studies have also reported inequalities in the uptake of cancer screening programmes among ethnic minorities [10-12]. Evidence from the cancer screening programmes in England and Scotland indicates lower participation among ethnic minorities, irrespective of socioeconomic background [13-15]. The presence and extent of ethnic inequalities in Wales is unknown. This information gap is contrary to the legal and policy commitments in the UK and specifically from Wales to tackle health inequalities and promote racial equality [16, 17].

The global SARS-CoV-2 pandemic saw unprecedented disruption to cancer screening in 2020, with national lockdowns and prioritisation of COVID-19 services causing many screening programmes to be paused [18, 19]. In Wales, the decision was taken to suspend invitations to the BSW programme from 20th March 2020 [20]. The suspension of invitations lasted approximately 4-months, with invitations recommencing in August 2020. Although this temporary suspension was necessary as referral for screening colonoscopy was not possible, the interruption of CRC screening programmes due to the pandemic impacted specialist referrals, diagnostic procedures, and treatment pathways [21]. In Wales, Greene and colleagues showed that CRC incidence decreased almost a fifth (17.2%) during 2020 overall, and by 59.9% during April 2020, compared to April 2019, coinciding with the strict lockdown implemented at the end of March 2020 [22]. Whilst there is national [22] and international evidence [23] for a rebound in uptake following the temporary suspension of the CRC programmes [24], the return is not expected to be even across the population, with concerns that underserved groups, including those socioeconomically disadvantaged, those considered clinically extremely vulnerable (CEV) and ethnic minorities fall behind due to the unequal impact that COVID-19 has had in these communities [23, 25].

As health systems recover from the disruption and reintroduce routine services, gaining a better understanding of the impact of COVID-19 on inequalities is crucial to inform future action to avoid further widening of inequalities in CRC screening and improve uptake in those subgroups of the population that may be slower to reengage. This retrospective register-based study aimed to examine the impact of the temporary suspension of the BSW programme and highlight inequalities that may benefit from tailored interventions. We first describe uptake patterns by sociodemographics for the first 3 months (August to October) of invitations following the programme’s suspension. Secondly, we compare uptake patterns in 2020/21 to the pre-pandemic period in 2019/20. Finally, to put the programme’s temporary suspension into a wider context, we explore uptake patterns of the same period from 2017/18 onwards.

## Methods

### Data preparation

All data accessed for this study was available within the Wales national trusted research environment (TRE) known as the Secure Anonymised Information Linkage (SAIL) Databank, hosted at Swansea University, Swansea, UK. All data acquisition into SAIL is completed through a standard split-file process, with anonymisation and encryption enabling anonymised individual-level, population-scale data to be accessed within SAIL, whereby an individual’s identity is removed and replaced with an Anonymised Linking Field (ALF), based on their NHS Number or combination of unique identifiers including name, date of birth, and sex [26, 27]. CRC screening data was extracted from the BSW dataset for all people aged 60-74 years of age and living in Wales at the time of their invitation. BSW data for 2020/21 was available from 1^st^ August 2020 (when the invitations re-started) to 30^th^ April 2021 (which was the most up-to-date BSW data at the time of the analysis). Therefore, we considered a 3-month invitation period (August to October 2020), with a maximum of 6-month follow-up period for participant response (to April 2021). To make comparisons possible, we selected the same period (invitations from August to October with a maximum 6 months follow-up) from the same period in the 3 preceding years (2019/20, 2018/19, and 2017/18) (Figure 1).

**Figure 1.**
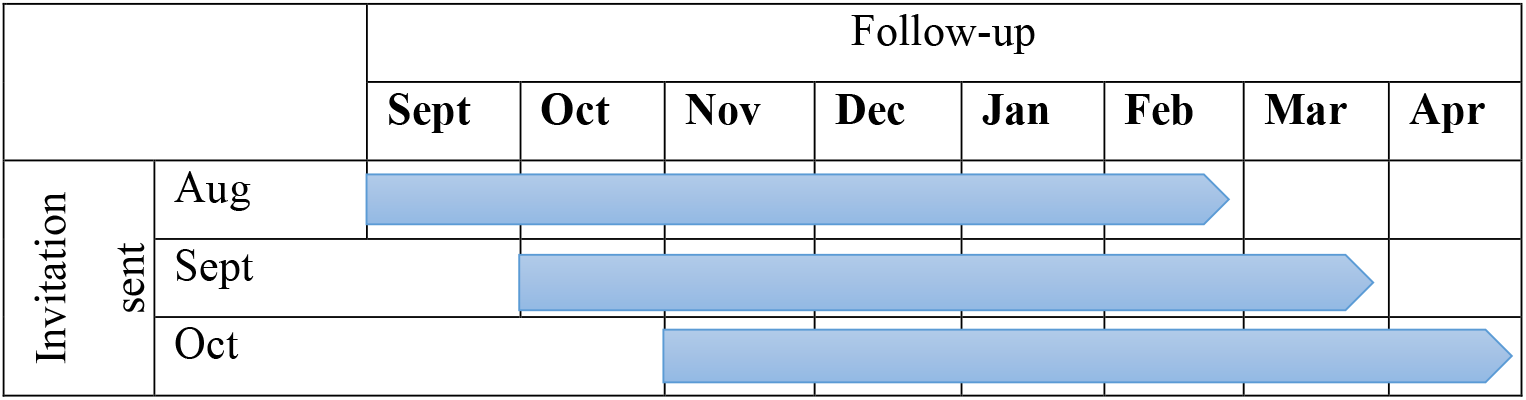
Waves of 3-month invitation period and 6-month follow-up period included in the analysis for each year.

Sex, age, and Lower-layer Super Output Area (LSOA) of residence were obtained from the Welsh Demographic Service Dataset (WDSD) as of 22^nd^ November 2021. LSOA of residence for individuals’ most recent address was linked to the Welsh Index of Multiple Deprivation (WIMD) 2019 income quintile to assign a measure of deprivation (28). The rural/urban classification of the individual’s residence was assigned by linking the LSOA to the Office for National Statistics (ONS) 2011 classification data [29]. Data from those individuals who were identified as CEV was sourced from the COVID-19 Shielded People List (CVSP) data, which is a list of all individuals in Wales who were identified as clinically extremely vulnerable to infection from COVID-19, based on clinician assessment and general practice records [30]. Finally, ethnic group was obtained from a linked data method available within SAIL using over 20 EHR and administrative data sources, including the ONS 2011 census, to harmonise the various values of defining ethnicity in each respective data source into a standardised ethnic group classification of the ONS five groups (white, mixed, Asian, black, and other) [31]. Repeated records, those with a low ALF matching rate (not allowing health data linkage), those outside the age range, non-routine recall invitations, and records from people deceased within 6-months of the invitation were excluded from the analysis (Figure 2).

**Figure 2.**
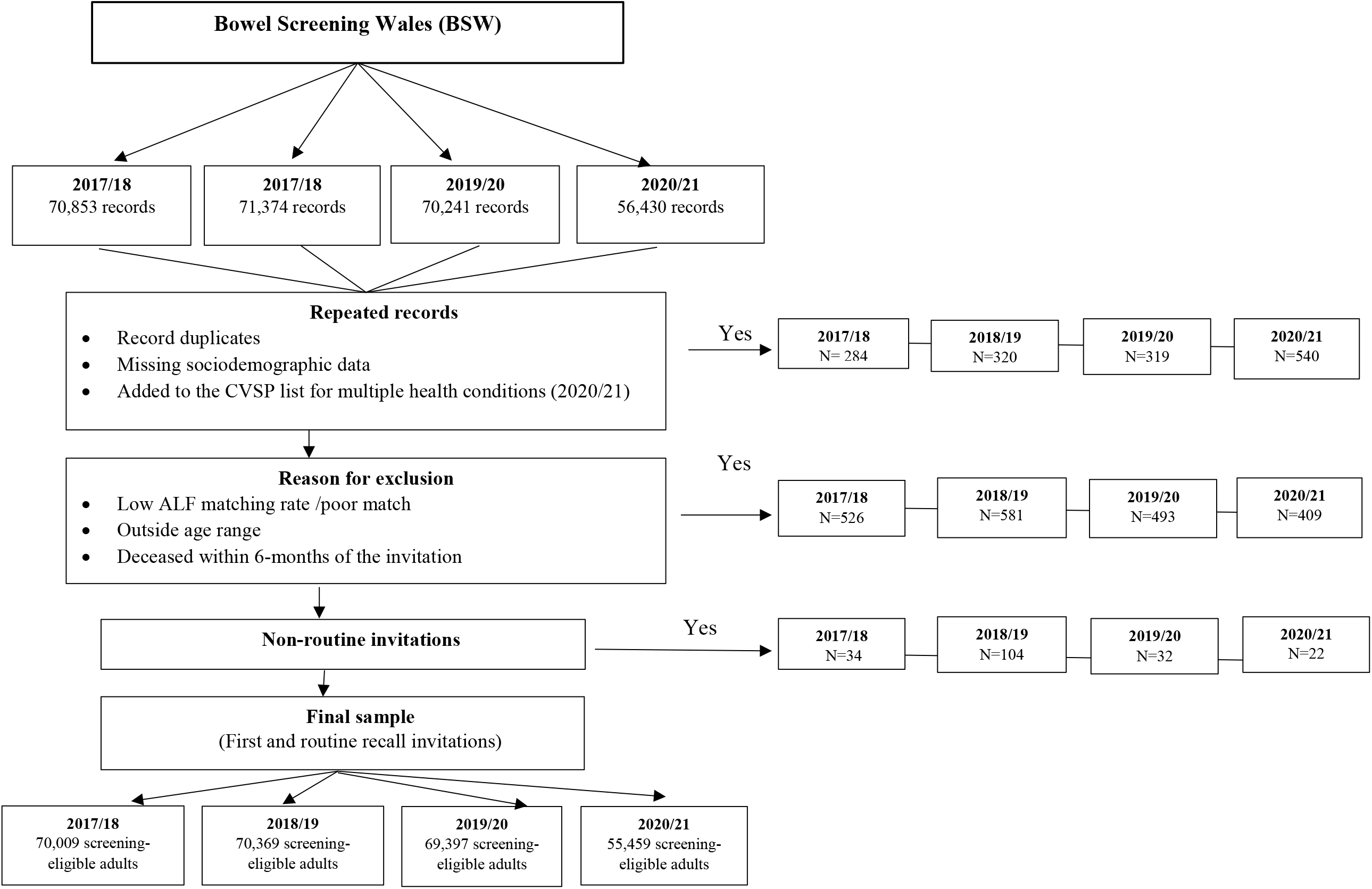
Data download

### Statistical analysis

Uptake was defined as the percentage of those invited who returned the test kit by post within 6-months of being sent the invitation letter. Descriptive statistics were calculated for all sociodemographic factors. Overall uptake percentages were calculated for each period of data, and adjusted proportions were calculated for sociodemographic subgroups using generalised linear models and reported as estimated marginal means with 95% confidence intervals. Binary logistic regression was used to examine differences in screening uptake in univariate analysis by ethnic group, age group, sex, income deprivation, and rural/urban location of residence, and in multivariate analysis adjusting for these factors. In addition, logistic regression models were conducted to compare uptake differences between periods. Statistical significance was set at p<0.05. Analysis was conducted with SPSS v.25 (IBM Corp., Armonk, New York, USA).

## Results

Table 1 shows the sample characteristics. The sample was predominantly of white ethnicity (83.0%), female (51.1%) and lived in rural locations (64.2%). In 4.8% of the overall sample, records could not be linked to LSOA data (Table 1).

**Table 1.**
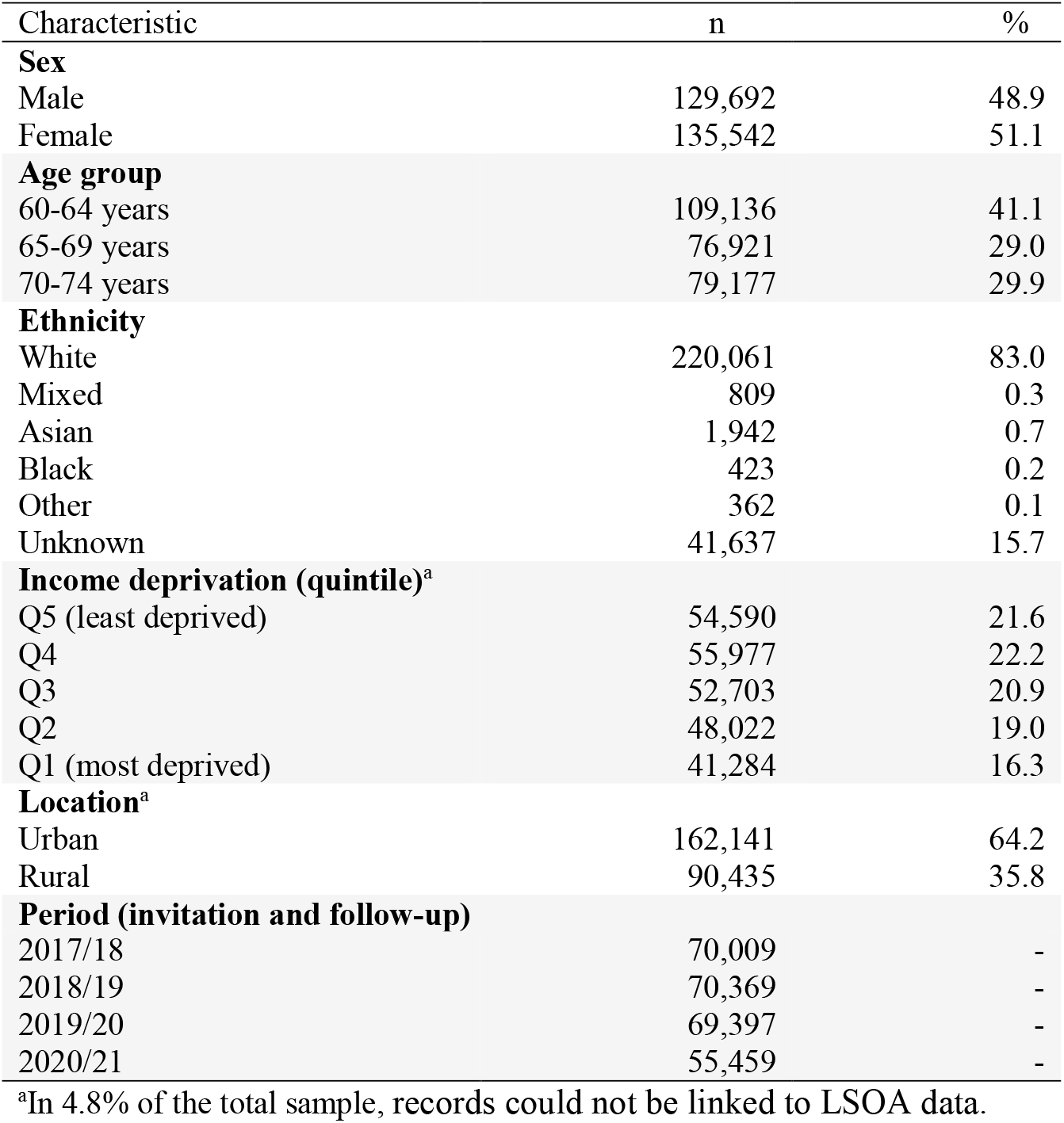
Sample characteristics (n=265,234)

### Overall uptake and variation by demographics following the temporary suspension of the programme (2020/21)

In 2020/21, overall uptake during the 3 months following the programme’s suspension (60.4%) was above the 60% Welsh standard (Figure 3). Uptake was higher in females (61.3% vs 59.5% for men; OR 1.07 95%, CI 1.04-1.11, p<0.001), older individuals, peaking in the group aged 70-74 years (68.5% vs. 55.0% for 60-64 years; OR 1.78, 95% CI 1.71-1.86) and in rural location (61.3% vs. 60.1% for urban location; OR 1.05, 95% CI 1.01-1.09) (Table 2; Figure 4a-4c).

**Figure 3.**
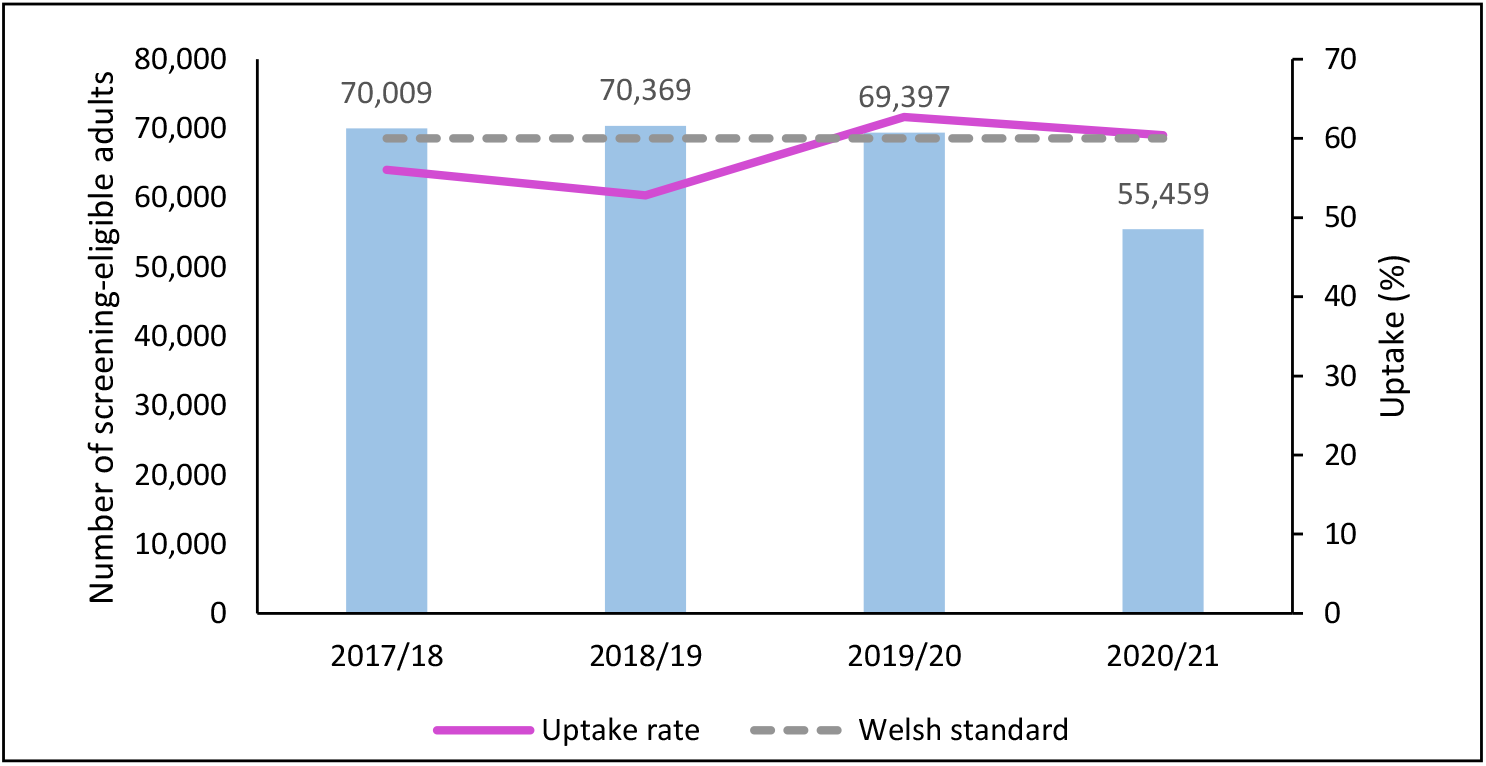
Number of screening-eligible adults and CRC uptake (%) by 3-month period

**Figure 4.**
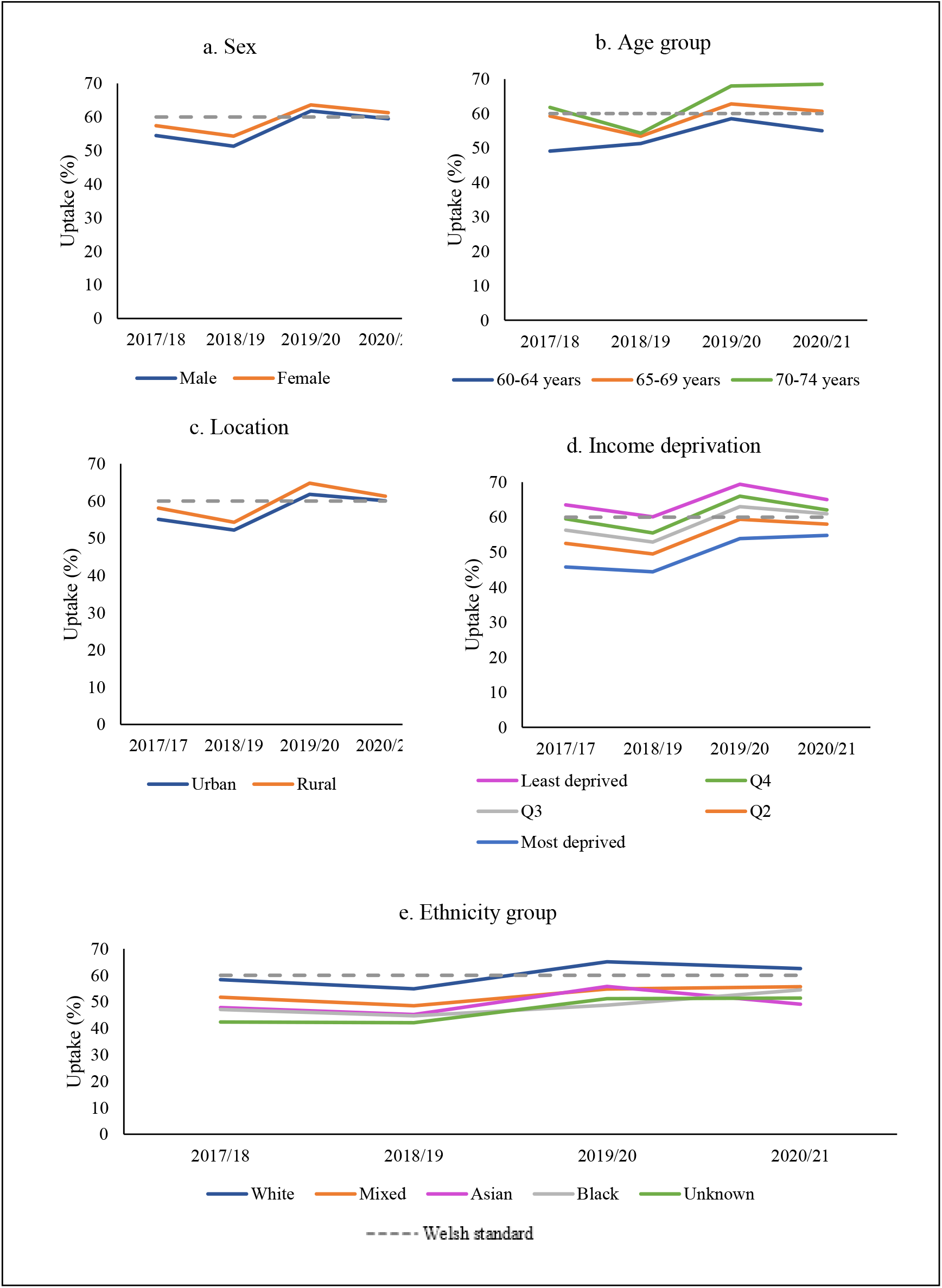
Uptake of CRC screening by 3-month period, stratified by demographics

**Table 2.** Univariable and multivariable analysis of BSW uptake for invitations during 3-month period in 2020/21 (Invitation period 1^st^ August-31^st^ October)

| Characteristic | Population (n) | Uptake (%) | OR | 95% CI | p | Adjusted uptake (%) | aOR | 95% CI | p |
| --- | --- | --- | --- | --- | --- | --- | --- | --- | --- |
| **Overall uptake** | 55,459 | 60.4 | - | - | - | 61.1 | - | - | - |
| **Sex** |  |  |  |  |  |  |  |  |  |
| Male | 27,537 | 59.5 | Reference |  |  | 60.0 | Reference |  |  |
| Female | 27,922 | 61.3 | 1.07 | 1.04-1.11 | **<0.001** | 61.1 | 1.05 | 1.01-1.09 | **0.011** |
| **Age group** |  |  |  |  |  |  |  |  |  |
| 60-64 years | 24,612 | 55.0 | Reference |  |  | 55.2 | Reference |  |  |
| 65-69 years | 14,923 | 60.7 | 1.26 | 1.21-1.32 | **<0.001** | 60.9 | 1.27 | 1.22-1.33 | **<0.001** |
| 70-74 years | 15,924 | 68.5 | 1.78 | 1.71-1.86 | **<0.001** | 68.3 | 1.76 | 1.69-1.84 | **<0.001** |
| **Location** |  |  |  |  |  |  |  |  |  |
| Urban | 18,723 | 60.1 | Reference |  |  | 60.5 | Reference |  |  |
| Rural | 33,644 | 61.3 | 1.05 | 1.01-1.09 | **0.009** | 60.5 | 0.99 | 0.96-1.04 | 0.848 |
| **Income deprivation** |  |  |  |  |  |  |  |  |  |
| Q5 (least deprived) | 11,297 | 65.0 | Reference |  |  | 64.5 | Reference |  |  |
| Q4 | 11,721 | 62.1 | 0.88 | 0.83-0.93 | **<0.001** | 62.0 | 0.90 | 0.85-0.95 | **<0.001** |
| Q3 | 10,968 | 61.0 | 0.84 | 0.80-0.89 | **<0.001** | 61.1 | 0.86 | 0.82-0.91 | **<0.001** |
| Q2 | 9,819 | 58.0 | 0.74 | 0.70-0.78 | **<0.001** | 58.1 | 0.76 | 0.72-0.80 | **<0.001** |
| Q1 (most deprived) | 8,562 | 54.8 | 0.65 | 0.62-0.69 | **<0.001** | 55.4 | 0.68 | 0.64-0.72 | **<0.001** |
| **Ethnic group** |  |  |  |  |  |  |  |  |  |
| White | 44,994 | 62.5 | Reference |  |  | 62.4 | Reference |  |  |
| Mixed | 194 | 55.7 | 0.75 | 0.57-1.00 | 0.051 | 59.0 | 0.86 | 0.64-1.16 | 0.331 |
| Asian | 401 | 49.1 | 0.58 | 0.48-0.71 | **<0.001** | 51.1 | 0.63 | 0.51-0.77 | **<0.001** |
| Black | 99 | 54.5 | 0.72 | 0.49-1.07 | 0.105 | 58.8 | 0.86 | 0.57-1.30 | 0.481 |
| Other | 68 | 51.5 | 0.64 | 0.40-1.03 | 0.063 | 53.3 | 0.69 | 0.42-1.12 | 0.132 |
| Unknown | 9,703 | 51.4 | 0.63 | 0.61-0.66 | **<0.001** | 51.5 | 0.64 | 0.61-0.67 | **<0.001** |
| **CEV status<sup>a</sup>** |  |  |  |  |  |  |  |  |  |
| Non-CEV | 262,129 | 60.7 | Reference |  |  | 57.5 | Reference |  |  |
| CEV | 5,296 | 57.8 | 0.89 | 0.84-0.94 | **<0.001** | 60.8 | 0.87 | 0.82-0.92 | **<0.001** |

<sup>a</sup>CEV=Clinical Extremely Vulnerable

During this period, uptake was socially graded between most and least income deprived quintiles, with those mid-income to least deprived (61.0% for Q3 to 65.0% for Q5) meeting the Welsh standard but not those in the most deprived income quintiles (58.0% for Q2 to 54.8% for Q1). Those from most income deprived areas were less likely (OR 0.65, 95% CI 0.62 to 0.692, p<0.001) to participate in the programme than those from least income deprived areas. Ethnic group comparisons showed that uptake in the Asian ethnic group (49.1%; OR 0.58, 95% CI 0.48-0.71, p<0.001) and those with unknown ethnicity (51.4%; OR 0.63, 95% 0.61-0.66, p<0.001) was lower than their white counterparts (62.5%) (Figure 4d-4e). Uptake was also significantly lower in people who were recommended to shield at the start of the pandemic (57.8% vs. 60.7%; OR 0.89, 95% CI 0.84-0.94, p<0.001) compared to non-CEV population. These differences remained significant after adjusting for all other sociodemographic variables, except that the difference between rural and urban locations was no longer significant (60.5% for both locations, p>0.05) (Table 2).

### Uptake compared to the pre-pandemic period (2019/20)

Overall, uptake decreased significantly during 2020/21 compared to the same 3-month period in 2019/20 (60.4% vs. 62.7%, OR 0.91, 95% CI 0.89-0.93, p<0.001) (Table 3, Figure 3). Uptake declined significantly for most sociodemographic groups (Table 3). However, when looking by age, no significant change was seen in those aged 70-74 years (68.5% vs. 68.0% for 2019/20, OR 1.03, 95% CI 0.98-1.07, p>0.241) and for area-level income-deprivation, there was no significant change for those in the most income deprived quintile (54.8% vs. 53.9% for 2019/20, OR 1.04, 95% 0.98-1.10, p=0.231). When looking by ethnic group, a significant decline was seen for those of white (62.5% vs. 65.1% for 2019/20, OR 0.89, 95% CI 0.87-0.92, p<0.001) and Asian ethnicity (49.1% vs. 55.8% for 2029/20, OR 0.77, 95% CI 0.59-0.96, p=0.045). This decline in uptake during 2020/21 among the Asian ethnic group was no longer significant after adjustment for other demographic variables, and there was no evidence of a statistically significant change in uptake for any other ethnicity groups (Table 3, Figure 4a-4e).

**Table 3.** BSW uptake variation by demographics between period in 2020/21 and period 2019/20 (Invitation period 1^st^ August-31^st^ October)

| Characteristic | Uptake (%)<br>2020/21 | Uptake (%)<br>2019/20 | OR | 95% CI | p | aOR<sup>a</sup> | 95% CI | p |
| --- | --- | --- | --- | --- | --- | --- | --- | --- |
| **Overall** | 60.4 | 62.7 | 0.91 | 0.89-0.93 | **<0.001** | 0.92 | 0.90-0.94 | **<0.001** |
| **Sex** |  |  |  |  |  |  |  |  |
| Male | 59.5 | 61.8 | 0.91 | 0.88-0.94 | **<0.001** | 0.92 | 0.89-0.95 | **<0.001** |
| Female | 61.3 | 63.6 | 0.90 | 0.99-0.93 | **<0.001** | 0.93 | 0.89-0.96 | **<0.001** |
| **Age group** |  |  |  |  |  |  |  |  |
| 60-64 years | 55.0 | 58.5 | 0.91 | 0.87-0.96 | **<0.001** | 0.86 | 0.83-0.89 | **<0.001** |
| 65-69 years | 60.7 | 62.8 | 0.91 | 0.87-0.95 | **<0.001** | 0.92 | 0.88-0.97 | **<0.001** |
| 70-74 years | 68.5 | 68.0 | 1.03 | 0.98-1.07 | 0.241 | 1.04 | 0.99-1.09 | 0.101 |
| **Location** |  |  |  |  |  |  |  |  |
| Urban | 60.1 | 61.8 | 0.93 | 0.91-0.96 | **<0.001** | 0.95 | 0.92-0.98 | **<0.001** |
| Rural | 61.3 | 64.8 | 0.86 | 0.82-0.89 | **<0.001** | 0.86 | 0.84-0.91 | **<0.001** |
| **Income deprivation** |  |  |  |  |  |  |  |  |
| Q5 (least deprived) | 65.0 | 69.4 | 0.82 | 0.77-0.86 | **<0.001** | 0.83 | 0.79-0.88 | **<0.001** |
| Q4 | 62.1 | 66.0 | 0.84 | 0.80-0.89 | **<0.001** | 0.86 | 0.82-0.91 | **<0.001** |
| Q3 | 61.0 | 63.0 | 0.92 | 0.87-0.97 | **0.001** | 0.94 | 0.89-0.99 | **0.022** |
| Q2 | 58.0 | 59.4 | 0.95 | 0.90-0.99 | **0.040** | 0.97 | 0.91-1.02 | 0.197 |
| Q1 (most deprived) | 54.8 | 53.9 | 1.04 | 0.98-1.10 | 0.231 | 1.06 | 0.99-1.12 | 0.153 |
| **Ethnic group** |  |  |  |  |  |  |  |  |
| White | 62.5 | 65.1 | 0.89 | 0.87-0.92 | **<0.001** | 0.91 | 0.88-0.93 | **<0.001** |
| Mixed | 55.7 | 54.8 | 1.04 | 0.70-1.54 | 0.854 | 1.25 | 0.82-1.90 | 0.297 |
| Asian | 49.1 | 55.8 | 0.77 | 0.59-0.96 | **0.045** | 0.82 | 0.63-1.08 | 0.168 |
| Black | 54.5 | 48.7 | 1.26 | 0.74-2.16 | 0.393 | 1.46 | 0.81-2.62 | 0.204 |
| Other | 51.5 | 55.4 | 0.85 | 0.46-1.58 | 0.611 | 0.96 | 0.48-1.92 | 0.911 |
| Unknown | 51.4 | 51.2 | 1.01 | 0.95-1.06 | 0.833 | 0.99 | 0.94-1.06 | 0.949 |

<sup>a</sup>2019/20 as the reference group

### Longer-term trends in CRC screening uptake

To put the temporary programme’s suspension during the 3-month period in 2020/21 into context, we examined differences in uptake during the same period from 2017/18 to 2020/21. Compared to period 2020/21, the uptake of the BSW programme was lower than the Welsh standard in 2018/19 (52.8%) and 2017/18 (56.0%) (Figure 3). When examining differences between sociodemographic groups, a similar pattern of inequalities was found across all years studied; with uptake in males, younger older adults (particularly those aged 60-64 years), those in the most income deprived quintiles (Q1-Q2), and ethnic minorities below the 60% Welsh standard (Supplementary tables S1-S3). However, in period 2019/20, uptake showed a significant increase among all sociodemographic groups, including low uptake groups such as males (61.8% vs. 51.3% for 2018/19; OR 0.65, 95% CI 0.63-0.67, p<0.001), those aged 60-64 years (58.5% vs. 51.3% for 2018/19; OR 0.75, 95% CI 0.72-0.77, p<0.001), those in the most income deprived quintile (53.9% vs. 44.4% for 2018/19; OR 0.69, 95% CI 0.65-0.72, p<0.001), and individuals of Asian (55.8% vs. 45.2% for 2018/19; OR 0.65, 95% CI 0.51-0.83, p<0.001), and unknown ethnic background (51.2% vs. 42.1% for 2018/19; OR 0.69, 95% CI 0.66-0.73). These differences remained significant after adjusting for other variables (Table 4, Figure 4a-4e).

**Table 4.**
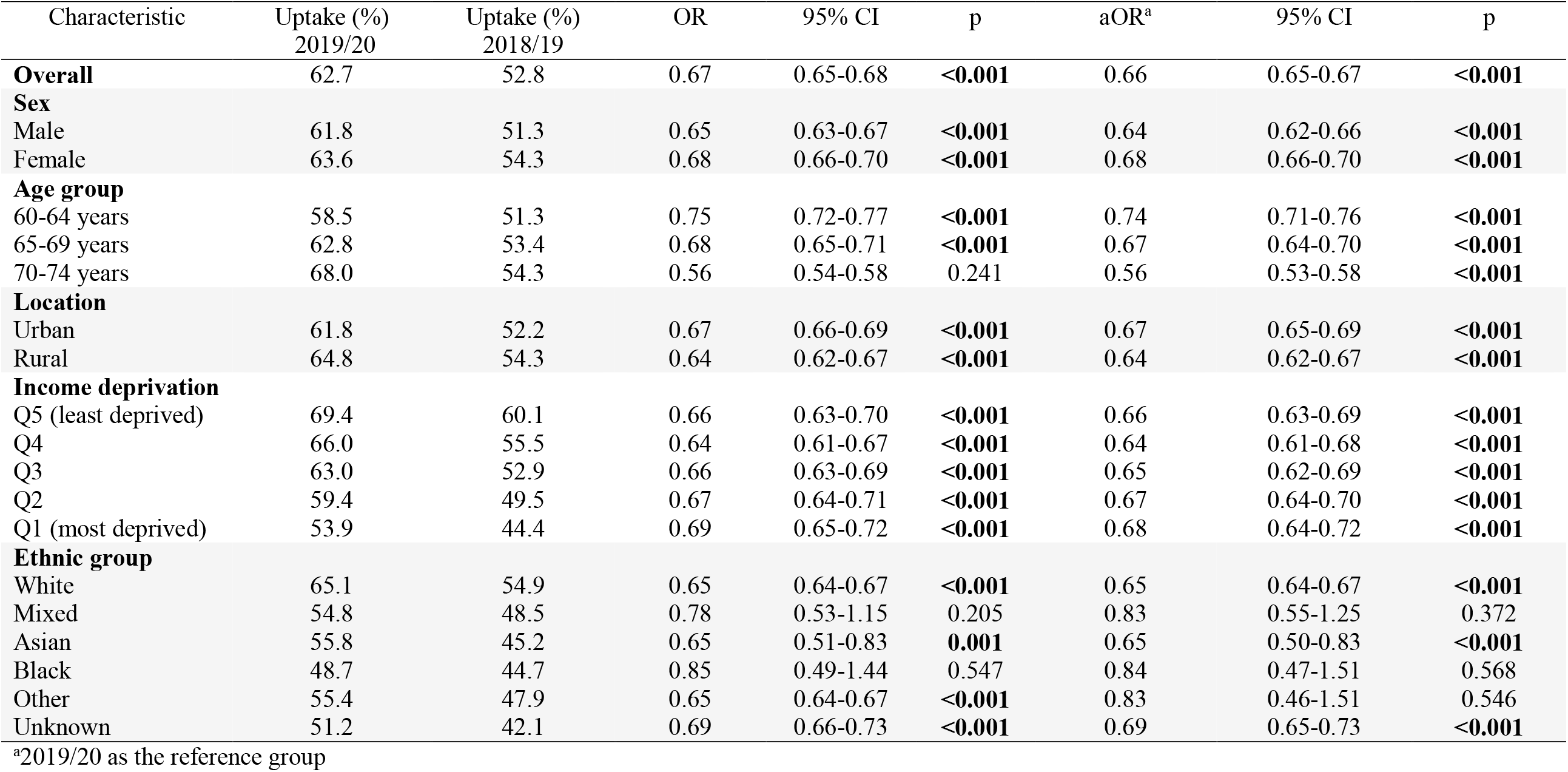
BSW uptake variation by demographics between period 2019/20 and period 2018/19 (Invitation period 1^st^ August-31^st^ October)

| Characteristic | Uptake (%)<br>2019/20 | Uptake (%)<br>2018/19 | OR | 95% CI | p | aOR<sup>a</sup> | 95% CI | p |
| --- | --- | --- | --- | --- | --- | --- | --- | --- |
| **Overall** | 62.7 | 52.8 | 0.67 | 0.65-0.68 | **<0.001** | 0.66 | 0.65-0.67 | **<0.001** |
| **Sex** |  |  |  |  |  |  |  |  |
| Male | 61.8 | 51.3 | 0.65 | 0.63-0.67 | **<0.001** | 0.64 | 0.62-0.66 | **<0.001** |
| Female | 63.6 | 54.3 | 0.68 | 0.66-0.70 | **<0.001** | 0.68 | 0.66-0.70 | **<0.001** |
| **Age group** |  |  |  |  |  |  |  |  |
| 60-64 years | 58.5 | 51.3 | 0.75 | 0.72-0.77 | **<0.001** | 0.74 | 0.71-0.76 | **<0.001** |
| 65-69 years | 62.8 | 53.4 | 0.68 | 0.65-0.71 | **<0.001** | 0.67 | 0.64-0.70 | **<0.001** |
| 70-74 years | 68.0 | 54.3 | 0.56 | 0.54-0.58 | 0.241 | 0.56 | 0.53-0.58 | **<0.001** |
| **Location** |  |  |  |  |  |  |  |  |
| Urban | 61.8 | 52.2 | 0.67 | 0.66-0.69 | **<0.001** | 0.67 | 0.65-0.69 | **<0.001** |
| Rural | 64.8 | 54.3 | 0.64 | 0.62-0.67 | **<0.001** | 0.64 | 0.62-0.67 | **<0.001** |
| **Income deprivation** |  |  |  |  |  |  |  |  |
| Q5 (least deprived) | 69.4 | 60.1 | 0.66 | 0.63-0.70 | **<0.001** | 0.66 | 0.63-0.69 | **<0.001** |
| Q4 | 66.0 | 55.5 | 0.64 | 0.61-0.67 | **<0.001** | 0.64 | 0.61-0.68 | **<0.001** |
| Q3 | 63.0 | 52.9 | 0.66 | 0.63-0.69 | **<0.001** | 0.65 | 0.62-0.69 | **<0.001** |
| Q2 | 59.4 | 49.5 | 0.67 | 0.64-0.71 | **<0.001** | 0.67 | 0.64-0.70 | **<0.001** |
| Q1 (most deprived) | 53.9 | 44.4 | 0.69 | 0.65-0.72 | **<0.001** | 0.68 | 0.64-0.72 | **<0.001** |
| **Ethnic group** |  |  |  |  |  |  |  |  |
| White | 65.1 | 54.9 | 0.65 | 0.64-0.67 | **<0.001** | 0.65 | 0.64-0.67 | **<0.001** |
| Mixed | 54.8 | 48.5 | 0.78 | 0.53-1.15 | 0.205 | 0.83 | 0.55-1.25 | 0.372 |
| Asian | 55.8 | 45.2 | 0.65 | 0.51-0.83 | **0.001** | 0.65 | 0.50-0.83 | **<0.001** |
| Black | 48.7 | 44.7 | 0.85 | 0.49-1.44 | 0.547 | 0.84 | 0.47-1.51 | 0.568 |
| Other | 55.4 | 47.9 | 0.65 | 0.64-0.67 | **<0.001** | 0.83 | 0.46-1.51 | 0.546 |
| Unknown | 51.2 | 42.1 | 0.69 | 0.66-0.73 | **<0.001** | 0.69 | 0.65-0.73 | **<0.001** |

<sup>a</sup>2019/20 as the reference group

## Discussion

This study provides population-level data on CRC screening in Wales and explores the impact of the programme’s temporary suspension due to COVID-19 on uptake inequalities following the reintroduction of the programme. It also compares patterns of inequalities with similar periods in the 3 preceding years.

Our findings suggest that the impact of the temporary suspension of the BSW programme on CRC screening uptake due to the COVID-19 pandemic was not large as overall uptake remained above the 60% Welsh standard [32]. These early findings are is in line with the early picture of the Scottish CRC programme [24]. However, the decline was significant, despite being a home-based test. It is possible that some people did not change the perceived risks of seeking diagnostic care due to the pandemic or had concerns over the capacity of the NHS. This decline is also important to consider in the context of the impact of the pandemic reported on other health services within the cancer pathway, including cancer-related referrals and diagnoses in Wales [22].

Comparing uptake rates between these 3-month periods in 2019/20 and 2020/21, some groups were less likely to engage with screening services. Engagement was lower than previously seen for younger older adults (<70 year old age groups, but particularly those aged 60-64 year olds) while those aged 70-74 year olds showed no significant decline. The drop in uptake in 2020/21 was larger in rural residents as well as in those living in the least deprived income areas (Q3 to Q5). This finding contrasts with no change in uptake among those living in more deprived income areas (Q1 to Q2), which is a positive finding. Nonetheless, levels of screening uptake for the most income deprived remained both below the Welsh standard and the levels seen for those living in the least income deprived areas. Uptake declined in both sexes but continued to be significantly higher in females. For ethnic minorities, small sample sizes precluded us from detecting statistically differences in uptake by ethnicity, particularly in the Asian and black ethnic groups such as those found in the English CRC screening programme [14,33]. Overall, this suggests that the impact of the programme’s temporary suspension on inequalities varied. Across different demographic characteristics, those less likely to engage with screening before the pandemic were not always those who showed the biggest reduction in uptake when screening services resumed.

Inequalities observed by sex, age, and deprivation were observed in all periods studied. These inequalities are similar to what has been reported internationally and in the English and Scottish CRC screening programmes since their inception [7,9,34]. In addition, our study supports findings from other studies conducted in the UK, indicating that ethnicity influences cancer screening, irrespective of income deprivation and rural/urban residence [12,13]. The mechanism driving this is beyond the scope of this study, but the large influence of cultural attitudes and beliefs relating to cancer screening well documented in the literature could explain the lower uptake seen in ethnic minorities [35, 36]. The full impact of the pandemic on CRC screening services amongst ethnic minorities requires further investigation.

Our study showed that overall uptake met the Welsh standard for the first time during the pre-pandemic period (2019/20), which could be related to the introduction of the easier-to-use test in September 2019 and continued efforts to target low uptake groups with different strategies such as repeat invitations [37]. Findings from the CRC screening programme in England also indicate that the introduction of the FIT resulted in higher uptake rates in males and across all deprivation quintiles [38]. We found that the difference between males and females started to reduce during 2019/20 due to greater gains in men, and uptake increased across all income quintiles but particularly in those in the most deprived income quintile. The new test’s introduction also appeared to positively impact all age and ethnic groups. However, our findings indicate that although there has been progress, uptake amongst low uptake groups including males, younger individuals, those in the most deprived quintiles, and ethnic minorities remains below the 60% Welsh standard.

## Strengths and limitations

To our knowledge this is the first study to include census linkage to look at CRC screening uptake by ethnicity, enabling a population-scale analysis of inequalities. Nonetheless, the ethnic grouping used in this study is broad and whilst grouping large visible ethnic groups is needed to avoid potentially disclosive numbers, it is problematic as it can hide key variations among ethnic groups. Furthermore, there are also important cultural and religious differences between ethnic minorities that have been shown to have an impact on cancer screening programmes [14, 35, 39]. Differences between white ethnic groups have also been found in other cancer screening programmes in the UK, but there is no data available for CRC screening programmes [13]. In addition, Wales is less ethnically diverse than most regions of England, with a small ethnically diverse population (5.9%) concentrated in urban locations such as Swansea, Cardiff, and Newport [40]. Given small numbers, comparisons between ethnic groups have limited statistical power. For the same reason, exploring the interaction between ethnic group and deprivation and/or location was not possible. BSW uptake in the population with an unknown ethnic group is low and this group needs further investigation. Improving the completeness and consistency of routinely collected ethnicity data in cancer screening programmes and greater transparency of linkage methods is crucial to obtaining disaggregated data by ethnic groups that can be used to plan public health strategies accordingly [41, 42].

We are only reporting on the first 3 months after the service resumed activities after the suspension of the service and comparing this to similar periods on previous years. The early findings from the BSW programme are encouraging; however, uptake will need to be continually monitored to understand if this changes over time. Analysing annual data with respect to ethnic groups is needed to identify any meaningful differences, but the required annual coverage of the data is not yet available in the SAIL Databank to analyse due to competing priorities for services as a result of the pandemic. Finally, the current study may be subject to ecological fallacy, where the associations may not be true at an individual level. However, all confounders included in our analysis are known to influence CRC screening participation.

Findings from this study add to the evidence base on inequalities in CRC screening and can inform future prioritisation strategies to promote equality of uptake and informed choice to assist with ongoing service recovery planning.

## Conclusions

Our findings are encouraging with overall uptake achieving the 60% Welsh standard during the first three months after the programme restarted in 2020 despite the disruption. The impact of the programme’s temporary suspension due to COVID-19 on inequalities varied. Across different demographic characteristics, those less likely to engage with screening prior to the pandemic were not always those who showed the biggest reduction in uptake following the reintroduction of the BSW programme. We showed that inequalities did not worsen during the first 3 months of invitations after the programme resumed activities but variations in CRC screening in Wales associated with sex, age, deprivation and ethnicity remain. This needs to be considered in targeting interventions to promote equality of uptake and informed choice to avoid exacerbating disparities in CRC outcomes as screening services recover from the pandemic.

## Supporting information

Supplementary material (Tables 1S-3S), and will be used for the link to the file on the preprint site.

STROBE checklist

ICMJE disclosure form

## Data Availability

The data used in this study are available in the SAIL Databank at Swansea University, Swansea, UK, but as restrictions apply they are not publicly available. All proposals to use SAIL data are subject to review by an independent Information Governance Review Panel (IGRP). Before any data can be accessed, approval must be given by the IGRP. The IGRP gives careful consideration to each project to ensure proper and appropriate use of SAIL data. When access has been granted, it is gained through a privacy protecting safe haven and remote access system referred to as the SAIL Gateway. SAIL has established an application process to be followed by anyone who would like to access data via SAIL at https://www.saildatabank.com/application-process.

## Declarations

## Acknowledgements

The authors would like to thank Dr Bethan Bowden and Dr Sikha de Souza for their comments on a previous version of the manuscript, and Laura Cowley and David Florentin for their support with data linkage. This study makes use of anonymised data held in the Secure Anonymised Information Linkage (SAIL) Databank. This work uses data provided by patients and collected by the NHS as part of their care and support. We would also like to acknowledge all data providers who make anonymised data available for research. We wish to acknowledge the collaborative partnership that enabled acquisition and access to the de-identified data, which led to this output. The collaboration was led by the Swansea University Health Data Research UK team under the direction of the Welsh Government Technical Advisory Cell (TAC) and includes the following groups and organisations: the SAIL Databank, Administrative Data Research (ADR) Wales, Digital Health and Care Wales (DHCW), Public Health Wales, NHS Shared Services Partnership (NWSSP) and the Welsh Ambulance Service Trust (WAST). All research conducted has been completed under the permission and approval of the SAIL independent Information Governance Review Panel (IGRP) project number 0911.

## Author’s contributions

D.B. was the main researcher and contributed to the design of the study, analysed the data and drafted the manuscript. A.A. completed the linked data method to create the ethnic groups. R.G. prepared or linked data sources to create the sample. S.H., J.S., D.H., G.G.,

A.D. and A.G. contributed to the design of this study and the revision of the manuscript. J.S. and K.H. contributed to the analysis plan, interpretation of the data, and manuscript revision. All authors read and approved the manuscript.

