## Supplementary material (Tables 1S-3S), and will be used for the link to the file on the preprint site. for "Inequalities in colorectal cancer screening uptake in Wales: an examination of the impact of the temporary suspension of the screening programme during the COVID-19 pandemic"

**Table 1S.** Univariable and multivariable analysis of BSW uptake for invitations during period 2019/20 (Invitation period 1st August-31st October)

| Characteristic | Population (n) | Uptake (%) | OR | 95% CI | p | Adjusted uptake (%) | aOR | 95% CI | p |
|---|---|---|---|---|---|---|---|---|---|
| **Overall** | 69,397 | 62.7 | - | - | - | 63.0 | - | - | **-** |
| **Sex** | | | | | | | | | |
| Male | 33,771 | 61.8 | Reference | | | 62.4 | Reference | | |
| Female | 35,626 | 63.6 | 0.92 | 0.90-0.95 | **<0.001** | 63.3 | 1.04 | 1.01-1.07 | **0.022** |
| **Age group** | | | | | | | | | |
| 60-64 years | 28,482 | 58.5 | Reference | | | 59.2 | Reference | | |
| 65-69 years | 18,532 | 62.8 | 1.96 | 1.15-1.24 | **<0.001** | 62.9 | 1.17 | 1.13-1.22 | **<0.001** |
| 70-74 years | 22,383 | 68.0 | 1.50 | 1.45-1.56 | **<0.001** | 67.4 | 1.44 | 1.38-1.49 | **<0.001** |
| **Location** | | | | | | | | | |
| Urban | 23,610 | 61.8 | Reference | | | 62.4 | Reference | | |
| Rural | 42,220 | 64.8 | 1.14 | 1.10-1.18 | **<0.001** | 63.7 | 1.09 | 1.05-1.12 | **<0.001** |
| **Income deprivation** | | | | | | | | | |
| Q5 (least deprived) | 14,380 | 69.4 | Reference | | | 69.0 | Reference | | |
| Q4 | 14,427 | 66.0 | 0.86 | 0.82-0.90 | **<0.001** | 65.7 | 0.86 | 0.81-0.90 | **<0.001** |
| Q3 | 13,723 | 63.0 | 0.75 | 0.72-0.79 | **<0.001** | 62.9 | 0.76 | 0.72-0.80 | **<0.001** |
| Q2 | 12,529 | 59.4 | 0.65 | 0.61-0.68 | **<0.001** | 59.6 | 0.66 | 0.62-0.69 | **<0.001** |
| Q1 (most deprived) | 10,771 | 53.9 | 0.52 | 0.49-0.54 | **<0.001** | 54.6 | 0.54 | 0.51-0.56 | **<0.001** |
| **Ethnic group** | | | | | | | | | |
| White | 57,281 | 65.1 | Reference | | | 65.0 | Reference | | |
| Mixed | 210 | 54.8 | 0.65 | 0.49-0.85 | **0.002** | 55.1 | 0.66 | 0.50-0.88 | **0.004** |
| Asian | 520 | 55.8 | 0.68 | 0.56-0.81 | **<0.001** | 56.7 | 0.71 | 0.60-0.85 | **<0.001** |
| Black | 117 | 48.7 | 0.51 | 0.35-0.73 | **<0.001** | 52.4 | 0.60 | 0.41-0.88 | **0.009** |
| Other | 101 | 55.4 | 0.67 | 0.45-0.99 | **0.044** | 55.8 | 0.68 | 0.45-1.02 | 0.062 |
| Unknown | 11,168 | 51.2 | 0.56 | 0.54-0.59 | **<0.001** | 51.7 | 0.57 | 0.55-60.0 | **<0.001** |

**Table 2S**. Univariable and multivariable analysis of BSW uptake for invitations during period 2018/19 (Invitation period 1st August-31st October)

| Characteristic | Population (n) | Uptake (%) | OR | 95% CI | p | Adjusted uptake (%) | aOR | 95% CI | p |
|---|---|---|---|---|---|---|---|---|---|
| **Overall** | 70,369 | 52.8 | - | - | **-** | 53.2 | - | - | **-** |
| **Sex** | | | | | | | | | |
| Male | 34,576 | 51.3 | Reference | | | 51.7 | Reference | | |
| Female | 35,793 | 54.3 | 0.89 | 0.86-0.91 | **<0.001** | 54.1 | 1.10 | 1.07-1.13 | **<0.001** |
| **Age group** | | | | | | | Reference | | |
| 60-64 years | 27,960 | 51.3 | Reference | | | 51.9 | 1.07 | 1.03-1.10 | **<0.001** |
| 65-69 years | 23,584 | 53.4 | 1.09 | 1.05-1.13 | | 53.4 | 1.08 | 1.04-1.12 | **<0.001** |
| 70-74 years | 18,825 | 54.3 | 1.13 | 1.08-1.17 | **<0.001** | 53.8 | 1.10 | 1.07-1.13 | **<0.001** |
| **Location** | | | | | | | | | |
| Urban | 23,881 | 52.2 | Reference | | | 52.8 | Reference | | |
| Rural | 43,254 | 54.3 | 1.09 | 1.06-1.13 | **<0.001** | 53.1 | 1.04 | 1.01-1.08 | **0.012** |
| **Income deprivation** | | | | | | | | | |
| Q5 (least deprived) | 14,190 | 60.1 | Reference | | | | Reference | | |
| Q4 | 14,843 | 55.5 | 0.83 | 0.79-0.87 | **<0.001** | 55.5 | 0.84 | 0.80-0.88 | **<0.001** |
| Q3 | 14,086 | 52.9 | 0.75 | 0.71-0.78 | **<0.001** | 52.8 | 0.75 | 0.71-0.79 | **<0.001** |
| Q2 | 12,804 | 49.5 | 0.65 | 0.62-0.68 | **<0.001** | 49.6 | 0.66 | 0.63-0.69 | **<0.001** |
| Q1 (most deprived) | 11,212 | 44.4 | 0.53 | 0.50-0.56 | **<0.001** | 44.7 | 0.53 | 0.51-0.56 | **<0.001** |
| **Ethnic group** | | | | | | | | | |
| White | 58,664 | 54.9 | Reference | | | 54.9 | Reference | | |
| Mixed | 198 | 48.5 | 0.77 | 0.59-1.02 | 0.072 | 50.3 | 0.83 | 0.62-1.11 | 0.207 |
| Asian | 538 | 45.2 | 0.58 | 0.57-80.3 | **<0.001** | 44.9 | 0.68 | 0.57-0.81 | **<0.001** |
| Black | 103 | 44.7 | 0.66 | 0.45-0.98 | **0.039** | 48.4 | 0.78 | 0.52-1.17 | 0.227 |
| Other | 94 | 47.9 | 0.76 | 0.50-1.13 | 0.174 | 50.0 | 0.82 | 0.54-1.24 | 0.335 |
| Unknown | 10,772 | 42.1 | 0.59 | 0.57-0.62 | **<0.001** | 42.3 | 0.60 | 0.58-0.63 | **<0.001** |

**Table 3S.** Univariable and multivariable analysis of BSW uptake for invitations during period 2017/18 (Invitation period 1st August-31st October)

| Characteristic | Population (n) | Uptake (%) | OR | 95% CI | p | Adjusted uptake (%) | aOR | 95% CI | p |
|---|---|---|---|---|---|---|---|---|---|
| **Overall** | 70,009 | 56.0 | - | - | **-** | 56.1 | - | - | **-** |
| **Sex** | | | | | | | | | |
| Male | 33,808 | 54.5 | Reference | | | 55.2 | Reference | | |
| Female | 36,201 | 57.4 | 0.89 | 0.86-0.92 | **<0.001** | 57.1 | 1.08 | 1.05-1.12 | **<0.001** |
| **Age group** | | | | | | | | | |
| 60-64 years | 28,082 | 49.1 | Reference | | | 49.9 | Reference | | |
| 65-69 years | 19,882 | 59.3 | 1.51 | 1.45-1.56 | **<0.001** | 59.3 | 1.47 | 1.42-1.53 | **<0.001** |
| 70-74 years | 22,045 | 61.8 | 1.67 | 1.61-1.73 | **<0.001** | 61.4 | 1.61 | 1.55-1.67 | **<0.001** |
| **Location** | | | | | | | | | |
| Urban | 24,221 | 55.1 | Reference | | | 55.9 | Reference | | |
| Rural | 43,023 | 58.1 | 1.13 | 1.10-1.17 | **<0.001** | 56.8 | 1.04 | 1.01-1.07 | **0.040** |
| **Income deprivation** | | | | | | | | | |
| Q5 (least deprived) | 14,723 | 63.5 | Reference | | | 63.1 | Reference | | |
| Q4 | 14,986 | 59.5 | 0.85 | 0.81-0.89 | **<0.001** | 59.3 | 0.85 | 0.81-0.89 | **<0.001** |
| Q3 | 13,926 | 56.3 | 0.74 | 0.71-0.79 | **<0.001** | 56.2 | 0.75 | 0.71-0.78 | **<0.001** |
| Q2 | 12,870 | 52.5 | 0.64 | 0.62-0.67 | **<0.001** | 52.7 | 0.65 | 0.62-0.68 | **<0.001** |
| Q1 (most deprived) | 10,739 | 45.8 | 0.49 | 0.46-0.51 | **<0.001** | 46.6 | 0.50 | 0.47-0.53 | **<0.001** |
| **Ethnic group** | | | | | | | | | |
| White | 59,122 | 58.4 | Reference | | | 58.3 | Reference | | |
| Mixed | 207 | 51.7 | 0.76 | 0.58-1.00 | 0.052 | 53.8 | 0.84 | 0.63-1.11 | 0.222 |
| Asian | 483 | 47.8 | 0.65 | 0.55-0.78 | **<0.001** | 49.9 | 0.72 | 0.60-0.86 | **<0.001** |
| Black | 104 | 47.1 | 0.64 | 0.43-0.93 | **0.021** | 51.3 | 0.78 | 0.52-1.16 | 0.214 |
| Other | 99 | 49.5 | 0.69 | 0.47-1.04 | 0.074 | 52.2 | 0.77 | 0.51-1.15 | 0.194 |
| Unknown | 9,994 | 42.4 | 0.53 | 0.50-0.55 | **<0.001** | 43.5 | 0.54 | 0.52-0.57 | **<0.001** |

### Declarations

#### Acknowledgements

The authors would like to thank Dr Bethan Bowden and Dr Sikha de Souza for their comments on a previous version of the manuscript, and Laura Cowley and David Florentin for their support with data linkage. This study makes use of anonymised data held in the Secure Anonymised Information Linkage (SAIL) Databank. This work uses data provided by patients and collected by the NHS as part of their care and support. We would also like to acknowledge all data providers who make anonymised data available for research. We wish to acknowledge the collaborative partnership that enabled acquisition and access to the de-identified data, which led to this output. The collaboration was led by the Swansea University Health Data Research UK team under the direction of the Welsh Government Technical Advisory Cell (TAC) and includes the following groups and organisations: the SAIL Databank, Administrative Data Research (ADR) Wales, Digital Health and Care Wales (DHCW), Public Health Wales, NHS Shared Services Partnership (NWSSP) and the Welsh Ambulance Service Trust (WAST). All research conducted has been completed under the permission and approval of the SAIL independent Information Governance Review Panel (IGRP) project number 0911.

#### Competing interests

The authors declare that they have no competing interests.

#### Author’s contributions

DB was the main researcher and contributed to the design of the study, analysed the data and drafted the manuscript. AA completed the linked data method to create the ethnic groups. RG prepared or linked data sources to create the sample. SH, JS, DH, GG, AD and AG contributed to the design of this study and the revision of the manuscript. JS and KH contributed to the analysis plan, interpretation of the data, and manuscript revision. All authors read and approved the manuscript.

#### Funding statement

This work was supported by the Con-COV team funded by the Medical Research Council (grant number: MR/V028367/1). This work was supported by Health Data Research UK, which receives its funding from HDR UK Ltd (HDR-9006) funded by the UK Medical Research Council, Engineering and Physical Sciences Research Council, Economic and

Social Research Council, Department of Health and Social Care (England), Chief Scientist Office of the Scottish Government Health and Social Care Directorates, Health and Social Care Research and Development Division (Welsh Government), Public Health Agency (Northern Ireland), British Heart Foundation (BHF) and the Wellcome Trust. This work was supported by the ADR Wales programme of work. The ADR Wales programme of work is aligned to the priority themes as identified in the Welsh Government's national strategy: Prosperity for All. ADR Wales brings together data science experts at Swansea University Medical School, staff from the Wales Institute of Social and Economic Research, Data and Methods (WISERD) at Cardiff University and specialist teams within the Welsh Government to develop new evidence which supports *Prosperity for All* by using the SAIL Databank at Swansea University, to link and analyse anonymised data. ADR Wales is part of the Economic and Social Research Council (part of UK Research and Innovation) funded ADR UK (grant ES/S007393/1). This work was supported by the Wales COVID-19 Evidence Centre, funded by Health and Care Research Wales.

**Availability of data and materials**

The data used in this study are available in the SAIL Databank at Swansea University, Swansea, UK, but as restrictions apply they are not publicly available. All proposals to use SAIL data are subject to review by an independent Information Governance Review Panel (IGRP). Before any data can be accessed, approval must be given by the IGRP. The IGRP gives careful consideration to each project to ensure proper and appropriate use of SAIL data. When access has been granted, it is gained through a privacy protecting safe haven and remote access system referred to as the SAIL Gateway. SAIL has established an application process to be followed by anyone who would like to access data via SAIL at https://www.saildatabank.com/application-process
